# Viruses such as SARS-CoV-2 can be partially shielded from UV radiation when in particles generated by sneezing or coughing: Numerical simulations

**DOI:** 10.1101/2020.11.19.20233437

**Authors:** David C. Doughty, Steven C. Hill, Daniel W. Mackowski

## Abstract

UV radiation can inactivate viruses such as SARS-CoV-2. However, designing effective UV germicidal irradiation (UVGI) systems can be difficult because the effects of dried respiratory droplets and other fomites on UV light intensities are poorly understood. Numerical modeling of UV intensities inside virus-containing particles on surfaces can increase understanding of these possible reductions in UV intensity. We model UV intensities within spherical approximations of virions randomly positioned within spherical particles. The model virions and dried particles have sizes and optical properties to approximate SARS-CoV-2 and dried particles formed from respiratory droplets, respectively. Wavelengths used are 260 nm (germicidal UVC) and 302 nm (solar UVB). In 5- and 9-μm diameter particles on a surface, illuminated by 260-nm UV light from a direction perpendicular to the surface, 10% and 18% (respectively) of simulated virions are exposed to intensities less than 1/100^th^ of intensities in individually exposed virions (i.e., they are partially shielded). Even for 302-nm light, where the absorption is small, 11% of virions in 9-µm particles have exposures 1/100^th^ those of individually exposed virions. Calculated results show that shielding of virions in a particle can be strongly reduced by illuminating a particle either from multiple widely separated incident directions, or by illuminating a particle rotating in air (because of turbulence, Brownian diffusion, etc.) for a time sufficient to rotate through all orientations with respect to the UV illumination. Because highly UV-reflective paints and surfaces can increase the angular ranges of illumination, they appear likely to be useful for reducing shielding of virions.

## 1. Introduction

Viruses such as influenza, measles, smallpox, SARS and some noroviruses can be transmitted via particles and droplets in the air. Improved methods to reduce the transmission of viruses are needed. Ultraviolet (UV) light can inactivate SARS-CoV-2 (Ratnesar-Shumate et al., 2020; Schuit et al., 2020; Simmons et al., 2020) and other viruses (Kowalski, 2009; Sagripanti and Lytle, 2011 and 2013; Tseng and Li, 2007; Kim and Kang, 2018; Sagripanti and Lytle, 2020), and thereby help reduce transmission rates (Anderson et al., 2018; Memarzadeh et al. 2010). Solar UV radiation (primarily UVB at 280-315 nm), can inactivate SARS-CoV-2 (Schuit et al., 2020; Ratnesar-Shumate et al., 2020) and other microbes near the Earth’s surface (Sagripanti and Lytle, 2007). UVC (200-280 nm) is common for germicidal applications (Garcia de Abajo et al., 2020; Rutala et al., 2013; Hakim et al., 2019; Bang et al., 2018; Kowalski, 2009; Kujundzic et al., 2007; Walker and Ko, 2007).

Viruses may occur within droplets expelled from the upper or lower respiratory tract (Morowska and Milton, 2020; Milton et al., 2013). Such droplets tend to lose water rapidly at relative humidity (RH) below 95%. At RH less than 50% they can dry into somewhat spherical particles (Vejerano and Marr, 2018). Such dried particles may land on surfaces and subsequently be transferred to hands, faces, etc. Some viruses including SARS-CoV-2 can remain pathogenic in aerosol particles, either in air (Schuit et al., 2020; Marr et al., 2019) or after landing on a surface (van Dormelan et al., 2020; Schuit et al., 2020; Ratnesar-Shumate et al., 2020; Park et al., 2015; Sagripanti et al., 2010). Viruses and other microbes on surfaces (bare or combined with other materials) may be aerosolized (suspended in air) (Lighthart et al., 1993; Joung et al., 2017; Girardin et al., 2016). If the material was initially deposited from air it may be reaerosolized (resuspended) (Fisher et al., 2012; Kesavan et al., 2017; Krauter and Biermann, 2007; Layshock et al., 2012; Paton et al., 2015; Qian and Peccia, 2014; Lighthart et al., 1993). The resulting airborne particles may be inhaled. Viruses on surfaces may also be transferred from surfaces to eyes, nose or mouth by touching with hands.

Some relevant surfaces for virus transmission include those in restaurants, restrooms, healthcare facilities, and on facemasks (Fisher et al., 2012; Woo et al., 2012) and other personal protective equipment. The relative importance of different routes of transmission depend upon the virus, and on factors such as airflows, RH and the behaviors of the relevant persons. In the case of SARS-CoV-2, although many questions remain unanswered, there is considerable interest in using UV to inactivate virions on surfaces (Ratnesar-Shumate et al., 2020), and in air (Garcia de Abajo et al., 2020; Nardell and Nathavitharana, 2020; Beggs and Vital, 2020; Hessling et al., 2020).

A microbe in a particle may be partially shielded from UV by other materials in the particle (Kesavan et al., 2014; Osman et al. 2008; Handler et al., 2015). This shielding may result in microbes remaining infectious in particles even when they are exposed to sufficient UV light to inactivate them if exposed individually. Such shielding can make germicidal UV more difficult in settings where microbes are in particles. An improved understanding of how virions in particles can be shielded from UV light could help in effectively using UV to deactivate SARS-CoV-2.

Optical phenomena affecting shielding of viruses from UV include the following (see Fig. 1). i) *Absorption* of UV causes the intensity to decrease as it travels into the particle and so reduces the intensity of UV reaching the virion. ii) *Refraction* of UV at a surface of a particle can direct light away from a virion; the importance of refraction depends upon the shape of the object, the angle of the incident UV light with respect to the orientation of the particle, and on the optical properties of the particle and the material outside the particle. iii) *Reflection* at the outer surface of a particle can reduce the UV entering or exiting a particle; in inhomogeneous particles it can occur at surfaces separating regions within the particle. iv) *Diffraction and scattering* can increase UV intensities in regions which may otherwise be dark, in a ray optics approximation, because of refraction and absorption.

**Figure 1:**
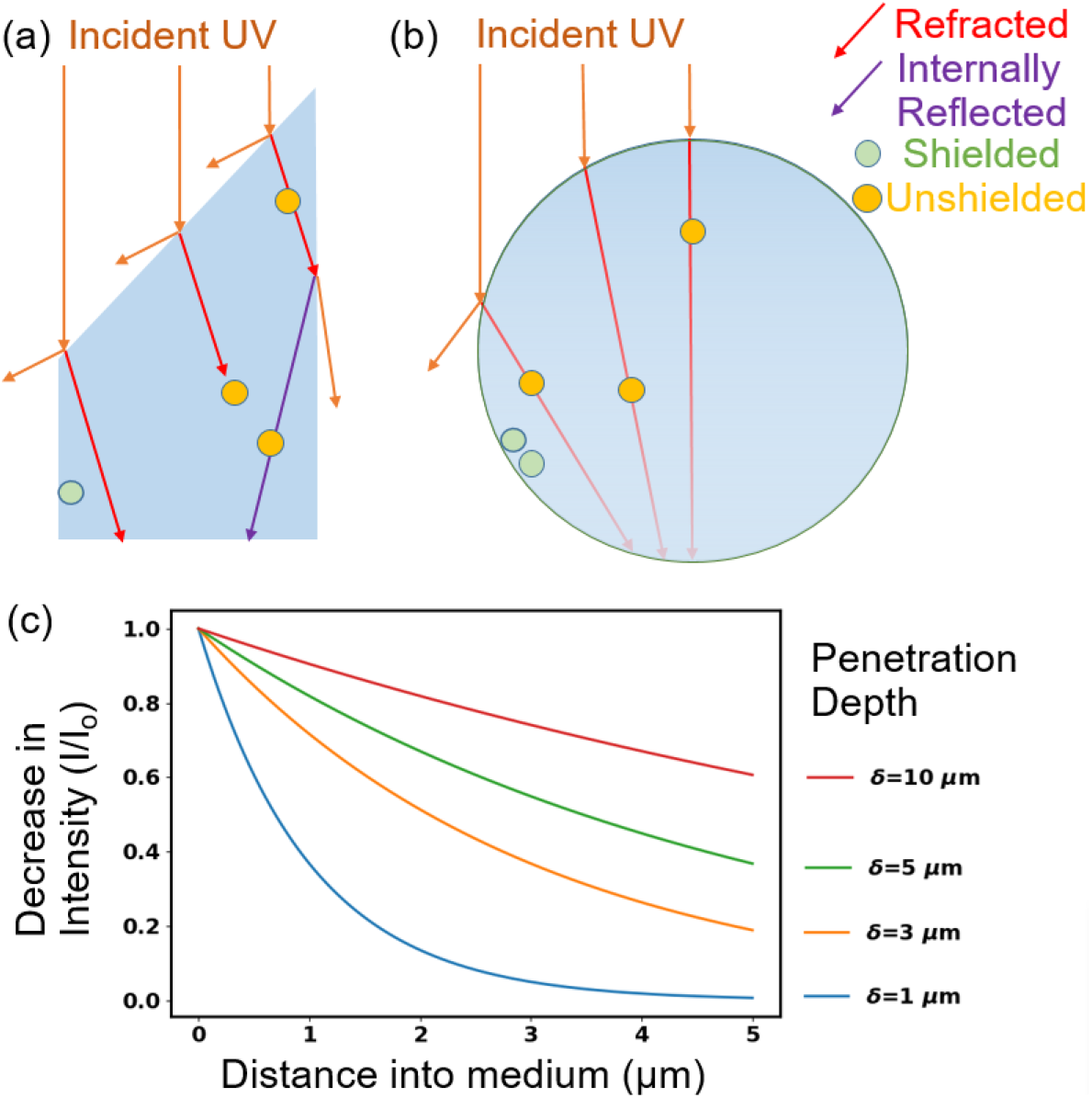
Illustration of optical features related to shielding of microbes such as viruses in particles from UV light. The material surrounding the particles is air. a) Refractive shielding in a non-spherical particle. b) Refractive and absorptive shielding in a spherical particle. If the virions are 100 nm diameter and the drawing is to scale, the large sphere is approximately 2 µm diameter. c) Decrease in intensity with distance into a medium as a function of penetration depth for δ similar to those used in this paper.

Models of the UV intensities in particles containing viruses can be useful in understanding UV inactivation and in improving UV decontamination systems. Valid models for the UV intensity within each virion can be combined with measured inactivation data to predict the numbers of virions likely to be inactivated within a particle. Validated models can predict results for large numbers of potential experiments that are impractical to perform in the laboratory or the field. Thus, such models can be essential tools for identification of decontamination techniques most appropriate for further testing. This paper focuses only on modeling of UV intensities within virions within particles, and not on modeling rates of inactivation.

Adequate methods to calculate the UV intensities within particles are required. Virions inside a dried saliva droplet resting on a surface and illuminated by a UV light source can be approximated as a number of homogeneous spherical virions distributed within a spherical particle (which is otherwise optically homogeneous) on a planar surface and illuminated by a UV plane wave. The UV intensities within the virions in the dried droplets in this model problem can be calculated using the Multi-Sphere T-Matrix (MSTM) method (Mackowski, 2008; Mackowski and Mishchenko, 2011, 2013). The MSTM provides an exact solution to the electromagnetic field equations (Maxwell’s equations) for problems which include non-overlapping spheres within spheres, and contact of one or more of the spheres with a planar surface.

In this paper we calculate the UV intensities inside of 100-nm diameter spherical particles to approximate virions which are inside a larger spherical particle of dried airway fluid. Each virion and larger particle has a complex refractive index (*m* = *m*_r_ + *i m*_i_) within the range of *m* estimated for SARS-CoV-2 and for dried particles of coughed, sneezed or exhaled droplets. Calculations of UV intensities within virions, and how virions in some locations are partially shielded from UV light at some positions inside the particle are done at two wavelengths (260 and 302 nm), for 100 nm virions to approximate SARS-CoV-2. Calculations are done to understand the amounts of shielding that could occur in dried droplets containing virions.

## 2. Methods

### 2.1 MSTM Calculations of UV intensities in Virions inside a particle on a surface or in air

Virions in this paper are approximated as homogenous spheres, typically inside a larger spherical particle which is in air or on a planar surface. In this paper the UV intensities in the spherical particles were calculated by solving the electromagnetic field equations (Maxwell’s equations) for a number of homogeneous spheres, separate or included within other spheres, and with the potential of being in contact with a flat surface, all illuminated by one or more plane waves (Mackowski, 2008; Mackowski and Mishchenko, 2011, 2013). In the MSTM as used here, a plane wave with irradiance *I*_o_ (or set of *n* plane waves with irradiance *I*_o_/*n* for each wave) illuminates either a naked virion or a spherical particle which contains a number of virions. The net rate of energy absorption by the *i*^th^ virion is *I*_o_π*a*_*i*_^2^*Q*_abs,*i*_, where *a*_*i*_ is the radius and *Q*_abs,*i*_ is the absorption efficiency, each for the *i*^th^ virion. The focus in this paper is on the absorption of UV energy by virions in the large particle relative to virions resting on (or very near, i.e., 10 nm) the surface. A main quantity illustrated here is the relative rate of absorption of UV energy of the *i*^th^ virion, *R*_*i*_ = Q_abs,i_ / Q_abs,single_ where Q_abs,single_ is the absorption efficiency of a virion resting on the same surface. Because the diameters and m for all virions in a given simulation are identical, *R*_*i*_ is the relative UV intensity within the *i*^th^ virion. The illumination used in the calculations is unpolarized because sunlight and UV lamps are unpolarized.

### 2.2 Optical Properties of Virions and Particles of Dried Respiratory Fluids

Droplets of airway fluids are assumed to be generated by sneezing or coughing. Their properties are assumed to be consistent with those of saliva or nasal fluids. The expelled droplets are assumed to dry into spherical particles; the dried particles of simulated saliva studied by Vejerano and Marr (2018) appeared to be somewhat spherical, at least for particles smaller than 25 µm. The dried particles modeled have diameters of 1, 5 and 9 μm. The dissolved solids in saliva are assumed to be 6 mg/cm^3^ (0.006×10^−12^ g/μm^3^), in the range of reported values (Edgar et al., 2012, Vejerano and Marr, 2018). We assume a density for the dry particle of 1.5 g/cm^3^ (1.5×10^−12^ g/μm^3^). For these values, droplets with diameters of 6.3, 31.5, and 56.7 μm, dry to particles with diameters of 1, 5 and 9 μm respectively.

Virions comprise at most a very small fraction of the mass of the solids in airway fluids. The concentrations of SARS-CoV-2 virions measured in COVID-19 patients and healthcare workers (Wyllie et al., 2020) ranged from 4×10^4^ to 4×10^10^ copies/mL in saliva and up to 5×10^9^ copies/mL in nasal fluids. The mass of a 100 nm diameter virion having an assumed density of 1.2 g/mL is 6×10^−13^ mg. The mass of 4×10^10^ of these virions (highest number measured in 1 mL by Wyllie et al. (2020)) is 0.024 mg, i.e., 1/240^th^ the mass of the other solids in the typical saliva assumed here. For the properties described in the previous paragraph, the maximum average numbers of virions per dried 9-μm particle is 3680 and in a dried 5-μm particle is 630. For almost all the other cases reported by Wyllie et al. (2020), the large majority of sneezed or coughed droplets and resulting dried particles would have no virions.

Each spherical particle is defined by a diameter (*d*) and a complex refractive index, *m* = *m*_r_+*im*_i_. The imaginary part of *m* is defined as *m*_i_ = λ/4π*δ*, where λ is the free-space wavelength, and δ is the penetration depth, a key parameter useful in understanding shielding by absorption. If *I(z)* is the intensity (*I*) of a UV planewave at distance *z* in a homogeneous medium, the fraction of light remaining at *z* is *I(z)/I*_0_ =*e*^-z/*δ*^, where the intensity at *z*=0 is *I*_0_, and *δ* is the distance at which the UV has decreased to 1/*e* (= 0.368) of *I*_0_. Also, *m*_i_ = λ*cε*/4π, where *c* is the concentration of the absorbing material and *ε* is the molar absorption coefficient (IUPAC 2014), also termed absorptivity (Hill et al. 2013; 2014; 2015), molar absorptivity, molar extinction coefficient or molar attenuation coefficient. The molar absorption coefficients of biological materials at the wavelengths used were estimated from the literature as described previously (Hill et al. 2013; Hill et al. 2014; Hill et al. 2015).

A representative complex refractive index of dried particles of respiratory fluids is estimated by first selecting representative concentrations in saliva and nasal fluids from the literature. The density of the dried solids is estimated as a weighted average of the densities of materials in the droplet. Using the initial concentrations and this density, the concentrations of the relevant materials in the dried droplet are calculated. The contribution of the *j*^th^ material to the total *m*_i_ of the dry particle, i.e., *m*_i,*j*_ is then is calculated using the concentration (g/g) and the extinction coefficient for the material as described previously (Hill et al., 2013, 2015). The total *m*_i_ is the sum of the *m*_i,j_. The concentrations of light-absorbing materials in dried droplets used here, as well as the *m*_i,j_, *m*_i_, *m*_r_ and penetration depths (δ) are given in Table 1. The main UV-light-absorbing materials in the 260 to 302 nm range in airway fluids are tryptophan and tyrosine (primarily in proteins), nucleic acids, and uric acid (Hawkins et al., 1963). At longer wavelengths the absorption by proteins, nucleic acids and uric acid tend to be so low that the penetration depths δ are many times larger than the diameter of most dried particles. Concentrations of components of saliva used here are selected to be representative of values reported by several sources (Edgar et al., 2012; Ben-aryeh et al., 1986; van Nieuw Amerongen et al., 2004; Veerman et al., 1996). The concentrations provided in Edgar et al. 2012 (Table 1.2, a compilation of 20 sources) were used for inorganic electrolytes (Na^+^, K^+^, Mg^2+^, Ca^2+^, Cl^-^, PO_4_^3-^, HCO_3_^-^, SCN^-^, and F^-^); nonabsorbing small molecules (glucose, lactate, lipids, urea, and ammonia); proteins and free amino acids; and the mucin glycoproteins. The concentration of protein used is 1.63 mg/ml. The combined concentration of mucins (MUC5B and MUC7) is 1.27 g/L, a concentration which includes both the amino acids and the carbohydrate of these glycoproteins. Typical mass fractions of airway mucin are 25% protein and 75% carbohydrate. Using these values, we calculated concentrations of carbohydrate in the mucins are 0.75 × 1.27 g/L = 0.95 g/L. Reported concentrations of DNA in saliva range from approximately 0.01 to 0.28 mg/ml (Poehls et al., 2018). The DNA concentration here is assumed here to be 0.12 mg/ml. Concentrations of RNA are assumed to be equal to those of DNA (0.12 mg/ml). We know of no reported measurements of non-viral RNA in saliva or nasal fluids. However, we assume that the DNA in these fluids arise from cells. Cells (human or bacterial) typically have several times as much RNA as DNA (Hill et al., 2013), and so the assumption of 0.12 mg/ml for RNA may be low. The concentration of uric acid used is 0.024 mg/ml, in the range of values measured (Hawkins et al. 1963; Araujo et al., 2020). Peden et al., (1990), demonstrate uric acid in nasal airway fluids. With the combination of values above, the total mass in the approximate airway fluid is approximately 5.77 g/L.

**Table 1:**
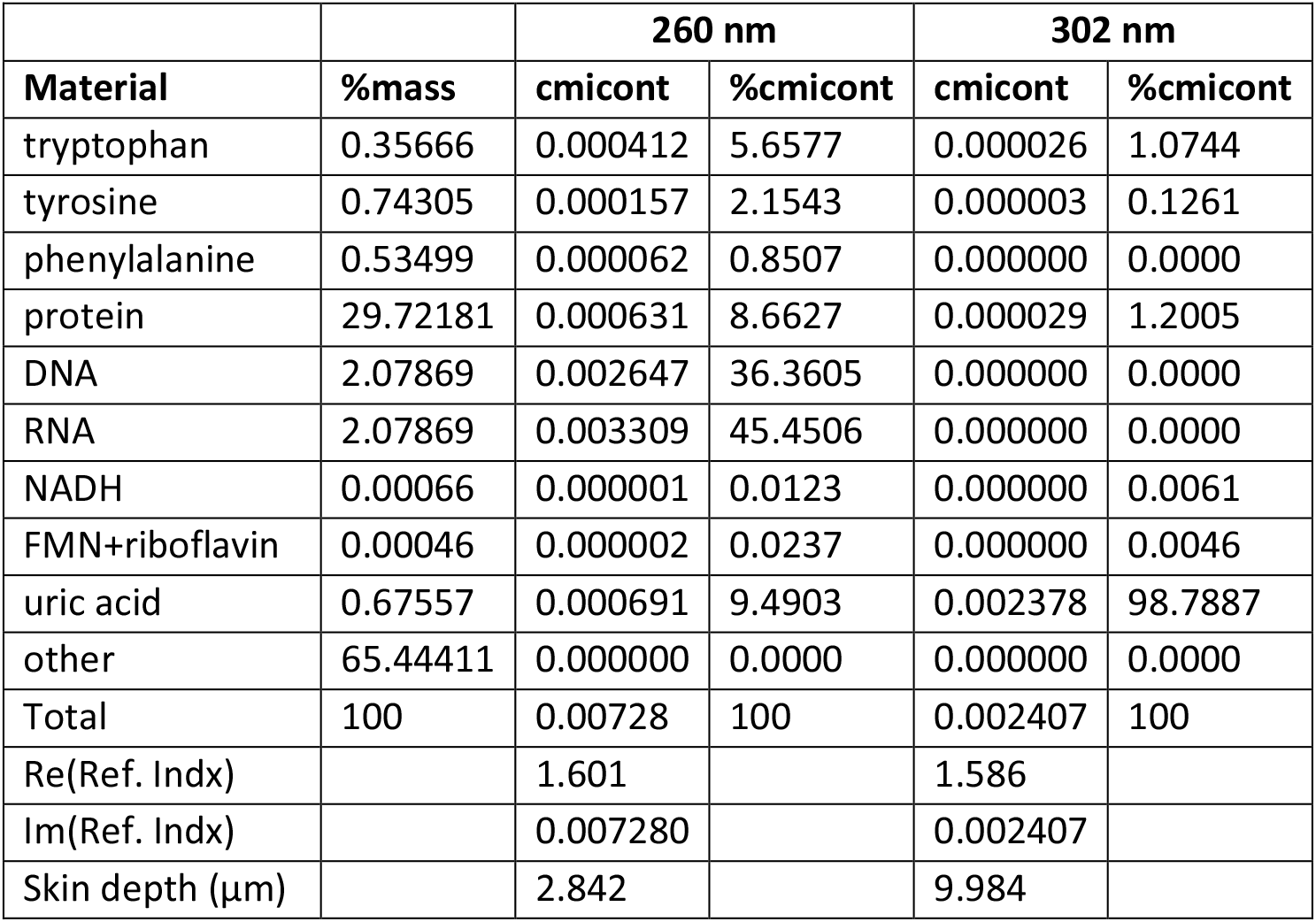
For dried droplets of respiratory fluids, the approximated concentrations (mass%) of the main UV absorbing materials and contributions to the imaginary part of the refractive index (total and percentage). The wavelengths used are in the top row. In the bottom three rows are the complex refractive index and the penetration (skin) depth.

|  |  | 260 nm |  | 302 nm |  |
| --- | --- | --- | --- | --- | --- |
| Material | %mass | cmicont | %cmicont | cmicont | %cmicont |
| tryptophan | 0.35666 | 0.000412 | 5.6577 | 0.000026 | 1.0744 |
| tyrosine | 0.74305 | 0.000157 | 2.1543 | 0.000003 | 0.1261 |
| phenylalanine | 0.53499 | 0.000062 | 0.8507 | 0.000000 | 0.0000 |
| protein | 29.72181 | 0.000631 | 8.6627 | 0.000029 | 1.2005 |
| DNA | 2.07869 | 0.002647 | 36.3605 | 0.000000 | 0.0000 |
| RNA | 2.07869 | 0.003309 | 45.4506 | 0.000000 | 0.0000 |
| NADH | 0.00066 | 0.000001 | 0.0123 | 0.000000 | 0.0061 |
| FMN+riboflavin | 0.00046 | 0.000002 | 0.0237 | 0.000000 | 0.0046 |
| uric acid | 0.67557 | 0.000691 | 9.4903 | 0.002378 | 98.7887 |
| other | 65.44411 | 0.000000 | 0.0000 | 0.000000 | 0.0000 |
| Total | 100 | 0.00728 | 100 | 0.002407 | 100 |
| Re(Ref. Indx) |  | 1.601 |  | 1.586 |  |
| Im(Ref. Indx) |  | 0.007280 |  | 0.002407 |  |
| Skin depth (μm) |  | 2.842 |  | 9.984 |  |

**Table 2:** For virions, the approximated concentrations (mass%) of the main UV-absorbing materials and contributions to the imaginary part of the refractive index (total and percentage). The wavelengths are in the top row. In the bottom three rows are the complex refractive index and penetration (skin) depth.

|  |  | 260 nm |  | 302 nm |  |
| --- | --- | --- | --- | --- | --- |
| Material | %mass | cmicont | %cmicont | cmicont | %cmicont |
| Tryptophan | 1.08750 | 0.001256 | 59.1656 | 0.000079 | 85.2596 |
| Tyrosine | 1.59500 | 0.000337 | 15.8599 | 0.000007 | 7.0469 |
| phenylalanine | 1.30500 | 0.000151 | 7.1173 | 0.000000 | 0.0000 |
| cystine | 0.36250 | 0.000029 | 1.3592 | 0.000002 | 2.4157 |
| protein | 72.50000 | 0.001772 | 83.5020 | 0.000088 | 94.7221 |
| RNA | 0.22000 | 0.000350 | 16.4980 | 0.000005 | 5.2779 |
| lipids | 27.20000 | 0.000000 | 0.0000 | 0.000000 | 0.0000 |
| other | 0.07999 | 0.000000 | 0.0000 | 0.000000 | 0.0000 |
| Total | 99.99999 | 0.002123 | 100 | 0.000092 | 100 |
| Re(Ref. Indx) |  | 1.59 |  | 1.575 |  |
| Im(Ref. Indx) |  | 0.002123 |  | 0.000092 |  |
| Skin depth ($\mu\text{m}$) $\delta$ | | 9.747 | | 259.852 | |

In estimating the complex refractive index of SARS-CoV-2, each virion was assumed to be 100 nm in diameter, where the outer 5 nm is a lipid bilayer. The virus genome, ssRNA (29,893 nucleotides, A=8954, T=9594, G=5863, U=5492) has a mass of 1.39 × 10^−17^ g, which is only 0.22% of the virion mass. The mass not in RNA or lipid is assumed to be protein, although we suspect other small molecules such as uric acid or NAD may be enclosed within the virion. The key parameter is the UV intensity in virions in a dried particle, relative to the UV intensity in an isolated virion resting on the same surface. In calculating the average m_v_, the average amino acid composition is taken as the average of all the amino acids specified by the genome. The different proteins are not weighted by their copy number or mass fraction mainly because we do not know the mass fractions to use. Each virion is assumed to be homogeneous. The code to calculate Table 1 and estimate optical properties for other combinations of primary metabolites is similar to that described previously (Hill et al. 2013; Hill et al. 2014; Hill et al. 2015).

The diameters (*d*) of dried respiratory particles in the calculations shown here are 1, 5 and 9 µm. The ratio *d*/δ can help in understanding the variations in calculated intensities. The intensity of an optical wave traveling a distance *z* in a medium with penetration depth δ is *I(z)/I*_0_ = *e*^*-z*/δ^. So, if *d*/δ = 5, then we may expect some positions inside the sphere to have intensities far smaller than the illuminating (incident) wave, maybe on the order of *e*^*-d*/δ^ = *e*^*-5*^ = 0.007 times. On the other hand, if *d*/δ is less than 0.3, the absorption in the sphere is relatively small, and significant protection from UV would need to arise from refractive effects. The location of virions within dried droplets of airway fluids has not been measured, so far as we know. The sensitivity of the distributions of the *R*_*i*_ to the numbers of virions in a dried particle was tested by calculating the *R*_*i*_ for particles with 4000, 3000, 2000, 1000, 500, 250, 100, 50, 25, and 10 virions. Effects of changes in virion numbers on the distribution of *R*_*i*_ calculated were below 0.1% until over 100 particles were used in a 9 µm dried droplet.

## 3. Results: UV Intensities in Virions in Particles of Dried Airway Fluids on a Surface

### 3.1 UV Intensities in Virions in particles illustrated in 3D

Figure 2 illustrates a 3D plot of 12,000 virions (calculated as 120 sets of 100 virions each) randomly positioned within a 9-μm sphere simulating a dried saliva particle illuminated with 260-nm UV light. The virions are color coded according to *R*_*i*_, the relative rate of absorption of UV energy by the *i*^th^ virion (relative to an isolated virion on the same surface). The dried particle is on a surface with *m=*1.4+*i* 0.0001, which has a relatively low reflectivity). The *m*_i_ and the *δ* are as shown in Table 1 for the dried saliva, and as in Table 2 for the virions. The incident wave is unpolarized; it propagates in a direction normal to the surface; and the large particle is spherical. Therefore, the intensity distribution would be independent of the angle around the sphere axis aligned with the direction of wave propagation. Also, the intensity is approximately independent of azimuthal angle for a sphere containing as many as 100 virions when the particle and virions have the optical properties used here. The lowest intensity regions, indicated by colors from blue to black, have UV absorption efficiencies that are over an order of magnitude smaller than the UV absorption of an individual virion on a surface. In these figures the shielded regions result from a combination of absorptive and refractive shielding.

**Figure 2:**
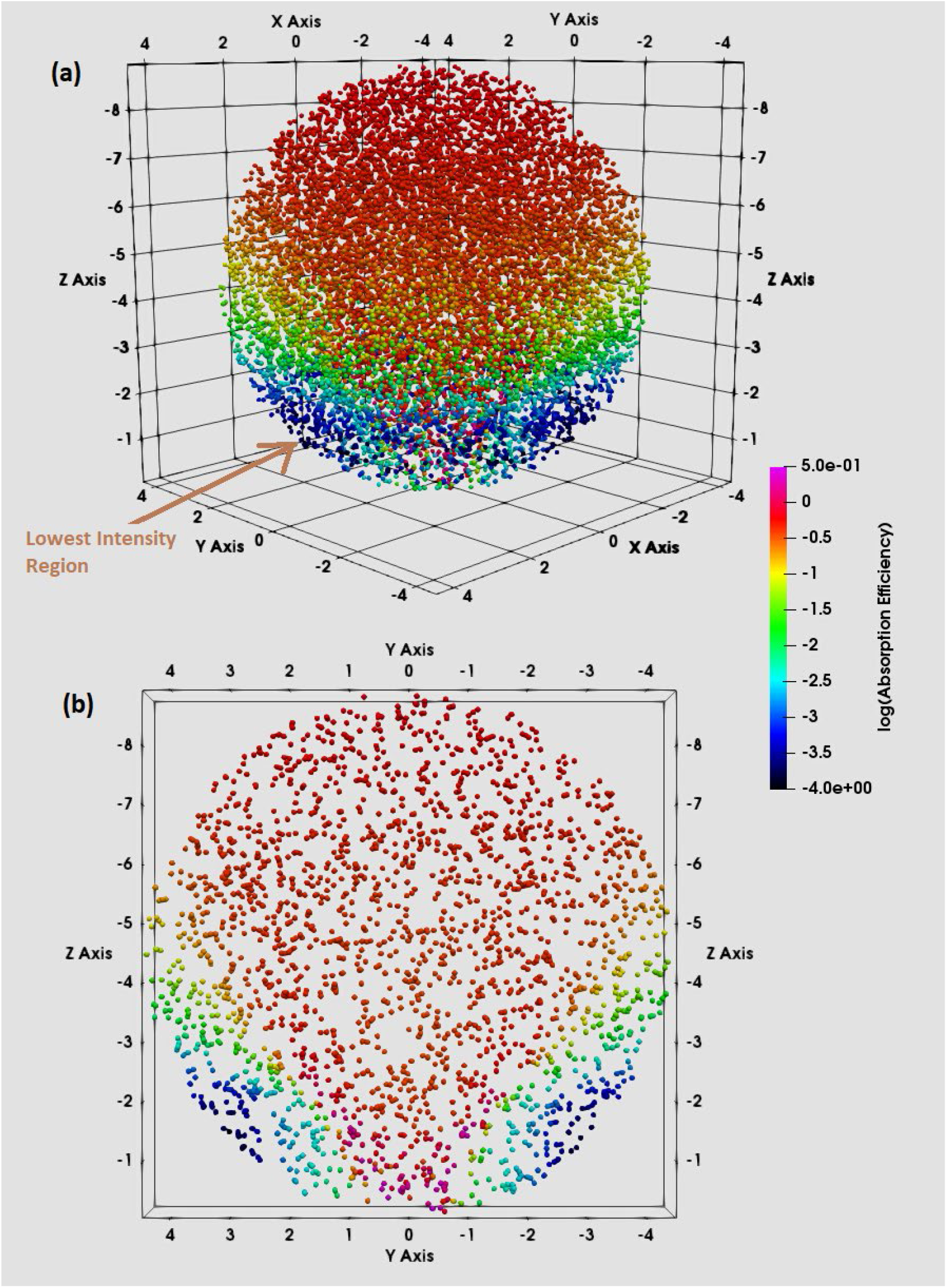
The spatial distribution of virions in 3D within a 9-µm diameter sphere approximating a spherical particle of dried airway fluids. The color of each virion indicates the R_i_, the relative UV intensity within that virion, and is proportional to the rate of absorption of UV energy by that virion. Results were calculated using the MSTM. The outer edge of the larger particle is not shown, but is just outside the outermost virion locations. The R_i_ were calculated using 120 sets of 100 virions each. B: A slice through the center of the sphere including all virions with -0.5 < x_center_ < 0.5 µm.

The highest intensities in Fig. 2 are at the top of the dried droplet and along an axis of the droplet in the direction of propagation of the wave (normal to the planar surface) which passes through the center of the sphere. This high-intensity region narrows into a cone of decreasing radius as it descends through the droplet and then partially reflects internally from the droplet surface as visualized using geometrical optics (Chowdhury et al., 1992). The most shielding from UV occurs in the outer edge of the particle on the side away from the incident light. Virions at the center of the particle are negligibly shielded by refraction from UV.

The 12,000 virions are used to illustrate the position-dependent variations in UV energy absorbed by the virions, not because a 9-μm particle is likely to carry such a large number of virions, but because a large number is needed to represent the distribution well. If 12,000 virions were in the particle simultaneously, virions with the optical properties used here would affect the UV intensities within other virions. Figure 2 may best be thought of as the distribution for an ensemble of 120 particles, each containing 100 virions.

### 3.2 Distributions of Virion Numbers vs UV Intensity for Different Penetration Depths

Figure 3 illustrates distributions of numbers of virions in different relative-UV-intensity bins vs relative-UV-intensity (log scale) for 9-µm particles shown in order of increasing *m*_*i*_. In all cases the particles are in air and the surface has *m* = 1.4+*i* 0.0001. In moving from top to bottom, the penetration depths decreases from 59.9 to 9.98 to 2.84 to 1.42 µm. Of these δ, the center two are as in Table 1 for 302 and 260 nm respectively. The δ in the top panel of Fig. 3 is six times the δ given in Table 1 for 302 nm. The bottom panel has a δ of ½ the δ estimated for 260 nm (in Table 1).

**Figure 3:**
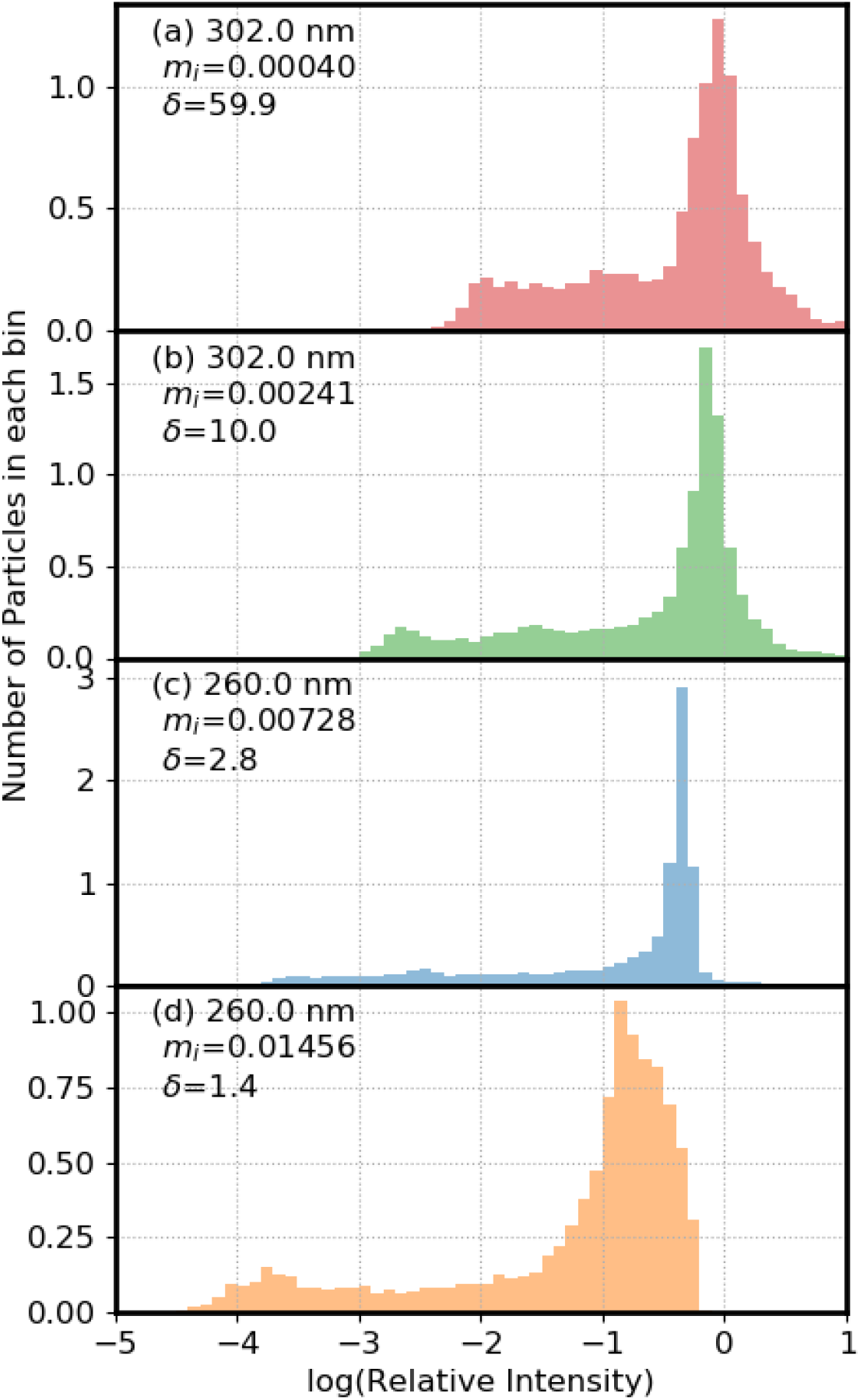
Relative numbers of virions in bins with the log(relative UV intensity) shown. The particles are 9-μm diameter and resting on a surface with m= 1.4 +i 0.0001. In all cases the number of virions (N) is 12000. a) m_i_ = 0.000401, δ=59.9 μm; b) m_i_ = 0.002407, δ=9.984 μm; c) m_i_ = 0.007280, δ=2.842 μm; d) m_i_ = 0.01456, δ=1.421 μm. The area under the histograms has been normalized to 1.

In moving from panel a) to d) the *R*_*i*_ of the bin with the highest number of virions decreases as follows, 0.89, 0.71, 0.45, and 0.14 (values for bin centers). In the case with the lowest *m*_*i*_ 59.5% of the virions have *R*_*i*_ greater than 0.8, and none have *R*_*i*_ < 0.001. As the *m*_*i*_ increases, the *R*_*i*_ decreases, and for the most absorbing case, 11.5% of virions have *R*_*i*_ < 0.001 and 1.9% have *R*_*i*_ < 0.0001 (Table 3).

**Table 3:** Percentage of virions below relative intensity of 1, 0.8, 0.5, 0.1, 0.01, 0.001, 0.0001. Wavelength (λ), m_i_, angle, orientation, and number of particles are shown. The wavelength is in units of nm, diameter is in units of microns. For orientation, “F” means Fixed, “All” means averaged over all orientations, “S” means on a surface, and “A” means in air. The percentages are shown in green in the table.

| Fig | $\lambda$<br>nm | d<br>μm | N | $m_i$ | angle<br>degrees | %$R_i < 1.0$ | %$R_i < 0.8$ | %$R_i < 0.5$ | %$R_i < 0.1$ | %$R_i < 0.01$ | %$R_i < 0.001$ | %$R_i < 0.0001$ |
| --- | --- | --- | --- | --- | --- | --- | --- | --- | --- | --- | --- | --- |
| 3a | 302 | 9 | 12000 | 0.0004 | $\beta=0, \alpha=0$ | 71.8 | 59.5 | 41.3 | 22.8 | 3.5 | 0 | 0 |
| 3b | 302 | 9 | 12000 | 0.00241 | $\beta=0, \alpha=0$ | 84.2 | 71.6 | 44.9 | 25.8 | 10.7 | 0.058 | 0 |
| 3c | 260 | 9 | 12000 | 0.00728 | $\beta=0, \alpha=0$ | 99.0 | 98.4 | 85.3 | 29.6 | 17.5 | 6.41 | 0 |
| 3d | 260 | 9 | 12000 | 0.01456 | $\beta=0, \alpha=0$ | 100 | 100 | 96.8 | 41.05 | 19.7 | 11.8 | 1.94 |
| 4a | 260 | 9 | 12000 | 0.00728 | $\beta=0, \alpha=0$ | 99.0 | 98.4 | 85.3 | 29.6 | 17.5 | 6.41 | 0 |
| 4b | 260 | 5 | 3000 | 0.00728 | $\beta=0, \alpha=0$ | 94.4 | 90.1 | 51.1 | 24.2 | 10.0 | 0 | 0 |
| 4c | 260 | 1 | 500 | 0.00728 | $\beta=0, \alpha=0$ | 68.2 | 52.2 | 20 | 0.2 | 0 | 0 | 0 |
| 5a | 260 | 9 | 12000 | 0.00728 | $\beta=0, \alpha=0$ | 99.0 | 98.4 | 85.3 | 29.6 | 17.5 | 6.41 | 0 |
| 5b | 260 | 9 | 12000 | 0.00728 | $\beta=40, \alpha=0$ | 98.9 | 98.5 | 83.0 | 28.7 | 14.7 | 4.7 | 0.275 |
| 5c | 260 | 9 | 12000 | 0.00728 | $\beta=80, \alpha=0$ | 88.1 | 78.3 | 63.9 | 30.1 | 15.8 | 6.07 | 0.958 |
| 6a | 260 | 9 | 12000 | 0.00728 | $\beta=0, \alpha=0$ | 99.0 | 98.4 | 85.3 | 29.6 | 17.5 | 6.41 | 0 |
| 6b | 260 | 9 | 12000 | 0.00728 | $\beta=0, 40, \alpha=0$ | 99.3 | 98.7 | 86.3 | 24.5 | 9.125 | 0.942 | 0 |
| 6c | 260 | 9 | 12000 | 0.00728 | $\beta=0, 80, \alpha=0$ | 98.7 | 95.6 | 70.7 | 18.7 | 6.82 | 1.43 | 0 |
| 6d | 260 | 9 | 12000 | 0.00728 | $\beta=80, \alpha=0, 180$ | 98.0 | 90.7 | 64.8 | 16.1 | 5.12 | 1.08 | 0 |
| 6e | 260 | 9 | 12000 | 0.00728 | $\beta=80, \alpha=0, 90, 180, 270$ | 99.0 | 93.8 | 65.7 | 5.675 | 0 | 0 | 0 |
| 6f | 260 | 9 | 12000 | 0.00728 | $\beta=0, \alpha=0 + \beta=80, \alpha=0, 180$ | 98.9 | 97.1 | 72.4 | 3.69 | 0 | 0 | 0 |
| 6g | 260 | 9 | 12000 | 0.00728 | $\beta=0, \alpha=0 + \beta=80, \alpha=0, 90, 180, 270$ | 99.8 | 97.5 | 70.8 | 1.425 | 0 | 0 | 0 |
| 7a | 260 | 5 | 3000 | 0.00728 | All | 100 | 70.7 | 56.2 | 0 | 0 | 0 | 0 |
| 7a | 260 | 5 | 3000 | 0.00728 | $\beta=0, \alpha=0$ | 91.9 | 84.3 | 45.8 | 22.8 | 8.6 | 0 | 0 |
| 7a | 260 | 5 | 3000 | 0.00728 | $\beta=0, \alpha=0$ | 94.4 | 90.1 | 51.1 | 24.2 | 10.0 | 0 | 0 |
| 7b | 260 | 9 | 4000 | 0.00728 | All | 100 | 100 | 90.9 | 0 | 0 | 0 | 0 |
| 7b | 260 | 9 | 12000 | 0.00728 | $\beta=0, \alpha=0$ | 98.6 | 98.1 | 72.4 | 28.7 | 16.6 | 6.68 | 0.0333 |
| 7b | 260 | 9 | 12000 | 0.00728 | $\beta=0, \alpha=0$ | 99.0 | 98.4 | 85.3 | 29.6 | 17.5 | 6.41 | 0 |

### 3.3 Distributions of Virion Numbers vs UV Intensity for Particle Diameters of 1, 5 and 9 µm

Figure 4 illustrates normalized distributions of numbers of virions within UV intensity bins for different sized particles resting on a surface and illuminated at 260 nm. The center-value *R*_*i*_ of the bins with the peak numbers occur at 0.45, 0.71, and 0.71, for panels a) to c) with particle diameters 9, 5 and 1 μm, respectively. In the case of the 1-µm particle, over 31% of the virions have *R*_*i*_ > 1.0, and 0.2% have *R*_*i*_ < 0.1. However, in the 9-µm case, 29.6% of virions have *R*_*i*_ < 0.1 and 6.4% have *R*_*i*_ < 0.001 (Table 3).

**Figure 4:**
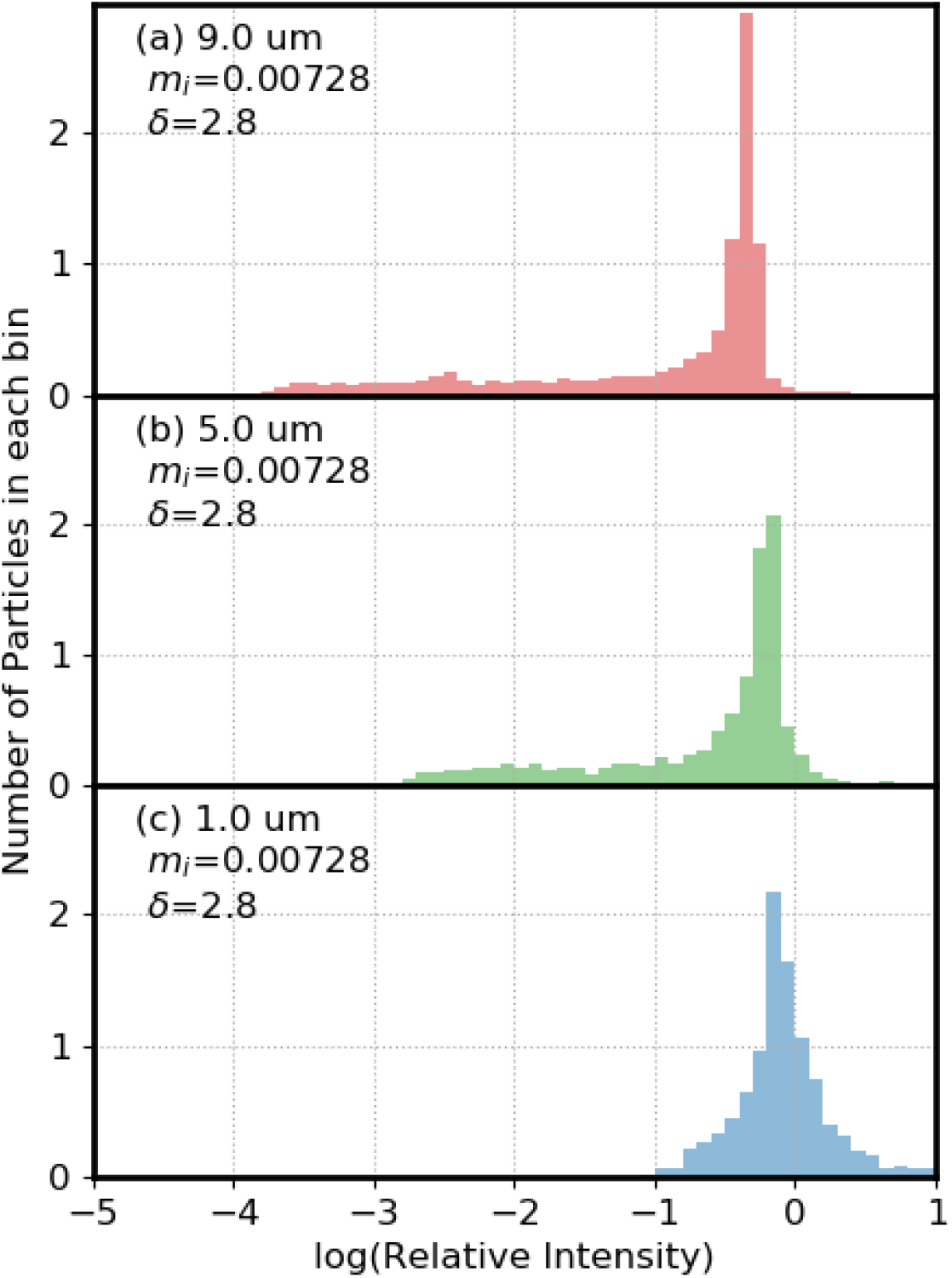
Relative numbers of virions in bins with the log(relative UV intensity) shown. Particle diameters are 9 μm in (a), 5 μm in (b), and 1 μm in (c). Particles are on a surface with m=1.4 + i 0.0001. The wavelength is 260 nm. The area under the histogram has been normalized to 1. The area under the histograms has been normalized to 1.

### 3.4 Histograms of UV Intensity with Illumination Waves at Different Angles from the Zenith

Figure 5 illustrates histograms for three incident angles (β) relative to the vertical, also termed the zenith angle. In Fig. 5a-c, β = 0, 40 and 80 degrees respectively. For these angles, the percentage of virions with *R*_*i*_ < 0.1 is approximately 30% (Table 3). The percentage of virions with *R*_*i*_ < 0.01 is approximately 15% (Table 3). The percentage of virions with *R*_*i*_ < 0.001 is approximately 5% (Table 3). However, although there are no virions with relative UV intensities below 0.0001 at an incident angle of zero degrees, at 80 degrees, approximately 1% of the virions had relative intensities below 0.0001. Also, when β = 80° there are a higher number of virions with *R*_*i*_ > 0.8 (Table 3).

**Figure 5:**
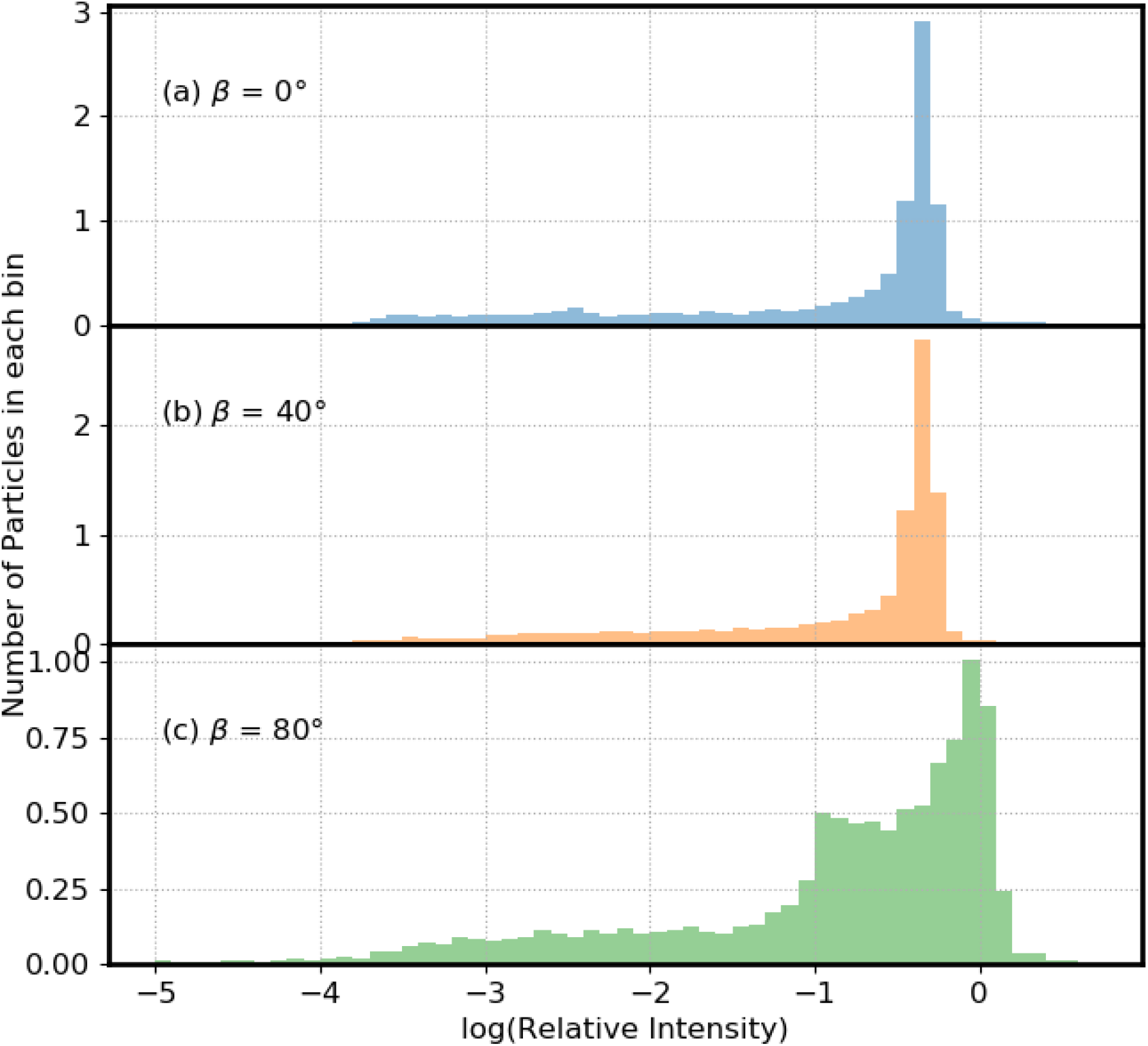
Relative numbers of virions in bins with the log(relative UV intensity) shown. Particle diameters are 9 μm. The wavelength of the incident UV light is 260 nm. Particles are on a surface with m=1.4 + i 0.0001. In (a) the UV light is incident perpendicular to the plane (blue). In (b) the UV light is incident from 40 degree from the normal (yellow). In (c) the incident UV is 80 degree from the normal (green). The area under the histograms has been normalized to 1.

### 3.5 Illumination from Multiple Angles Sequentially or Simultaneously

Figure 6 illustrates distributions of UV intensity in virions within a 9-μm particle on a surface illuminated by one-to five-planewaves. The illumination could be simultaneous or sequential: the resulting total UV intensities within virions are identical because the UV light source is not coherent. In Fig. 6a the illumination is from above the sphere (β =0). In Fig. 6b the angles (β, α) are (0, 0) and (40, 0) (the azimuthal angle is α). In Fig. 6c the two incident waves are defined by angles (0, 0) and (80, 0). In Fig. 6d the two incident waves are given by (80, 0) and (80, 180). In each of these four cases ((a) to (d)), some virions have *R*_*i*_ < 10^−3^ (Table 3).

**Figure 6:**
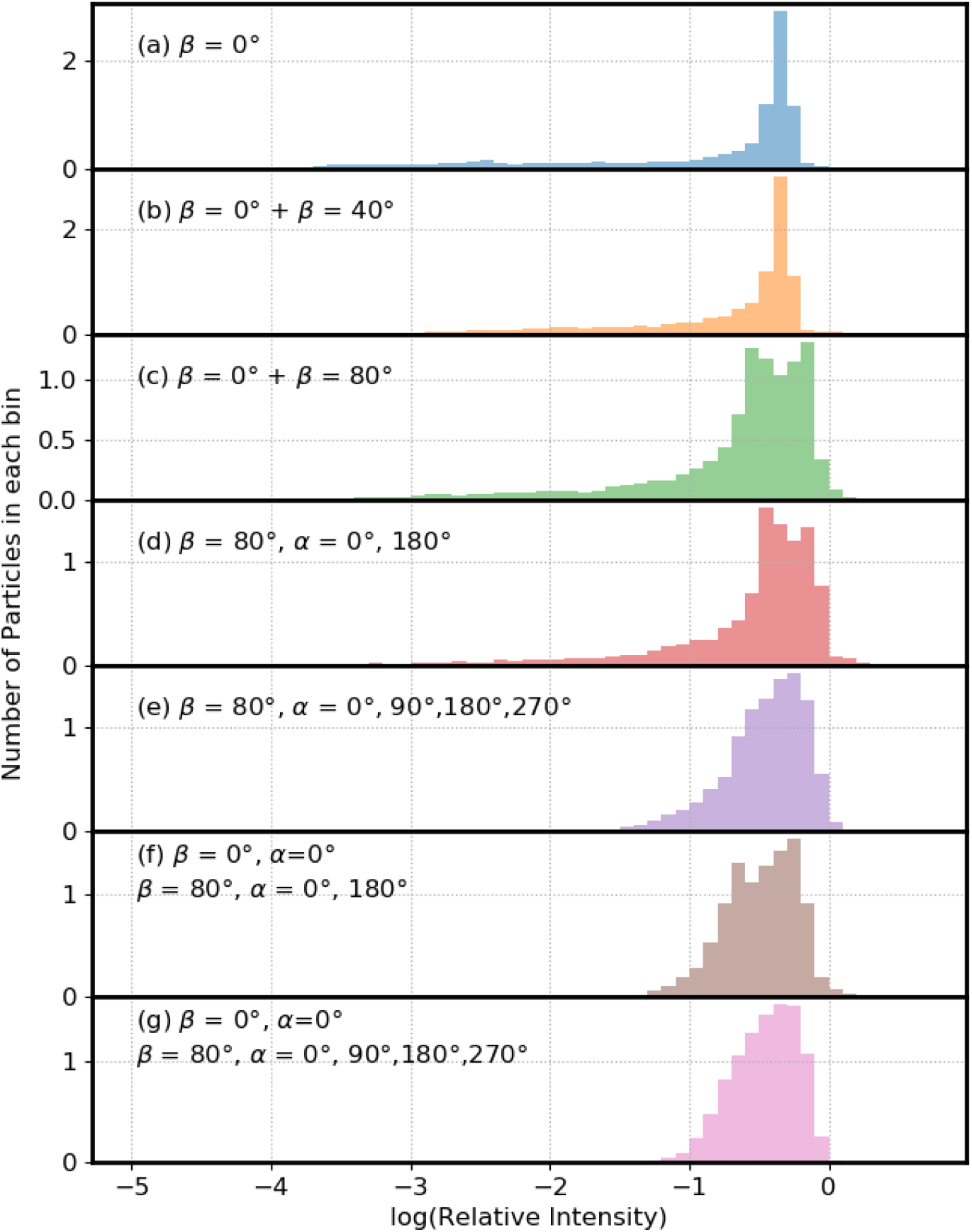
Relative numbers of virions in bins with the log(relative UV intensity) shown. Particle diameters are 9 μm. The wavelength of the incident UV light is 260 nm. Particles are on a surface with m=1.4 + i 0.0001. In (a) the UV light is incident perpendicular to the surface (blue). In (b) one UV wave is normal to the surface and the other is 40 deg from the normal. In (c) one UV wave is normal to the surface and the other is 80 deg from the normal. In (d) both waves are incident 80 deg from the normal to the surface but the azimuthal angle is 0 deg in one and 180 deg in the other. In (e) four waves, all incident at 80 degree from the normal, are used. They differ in having azimuthal angles of 0, 90, 180 and 270°. In (f), three incident UV waves are used, the two waves as in (d) but also with the normal-incidence wave of (a). In (g), five incident UV waves are used: the four waves as in (g) but also with the normal-incidence wave of (a). Each incident UV plane wave has the same intensity. To compute the intensities within virions the intensity for each angle of incidence is averaged. The area under the histograms has been normalized to 1.

In each of the last three panels, three or more incident UV waves are used and the minimum *R*_*i*_ increase significantly. In Fig. 6e four UV waves, incident from angles (80, 0), (80, 90), (80, 180) and (80,270) are used, and 5.7% of virions have *R*_*i*_ < 0.1. In Fig. 6f, three incident waves are used: two planewaves propagating primarily toward each other (at angles (80, 0) and (80, 180)), and the third wave traveling perpendicular to the surface. In this case, 3.7% of particles have *R*_*i*_ < 0.1, and the minimum *R*_*i*_ is 0.04. In Fig. 6g the particle is illuminated by five planewaves: one is from the vertical; the other four have β=80° and azimuthal angles = 0, 90, 180 and 270°. That is, the angles of the five waves are: (0, 0), (80, 0), (80, 90), (80, 180), and (80, 270). In this case, 1.4% of the particles have *R*_*i*_ < 0.1, and the minimum *R*_*i*_ is approximately 0.05.

For an objective of illuminating every virion in such particles with at least some desired UV intensity *I*_*o*_, these results suggest that illuminating with multiple waves as illustrated in Figs. 6e, and especially 6f and g, could provide a way to achieve that goal with much lower total UV than needed for illumination with one or two beams.

### 3.6 Particles in Air Illuminated from All Directions

In Fig. 7, the histograms with solid-color bars show the distributions of relative-UV-intensities in virions within 5- and 9-μm diameter particles. The particles are averaged over all orientations with respect to an incident UV planewave, calculated as described by Mackowski and Mishchenko (1996, 2011), which is equivalent to illuminating particles with equal-intensity UV-light from all directions. Also shown in Fig. 7 are the UV intensity histograms for the fixed-orientation cases (histograms made with thin lines).

**Figure 7:**
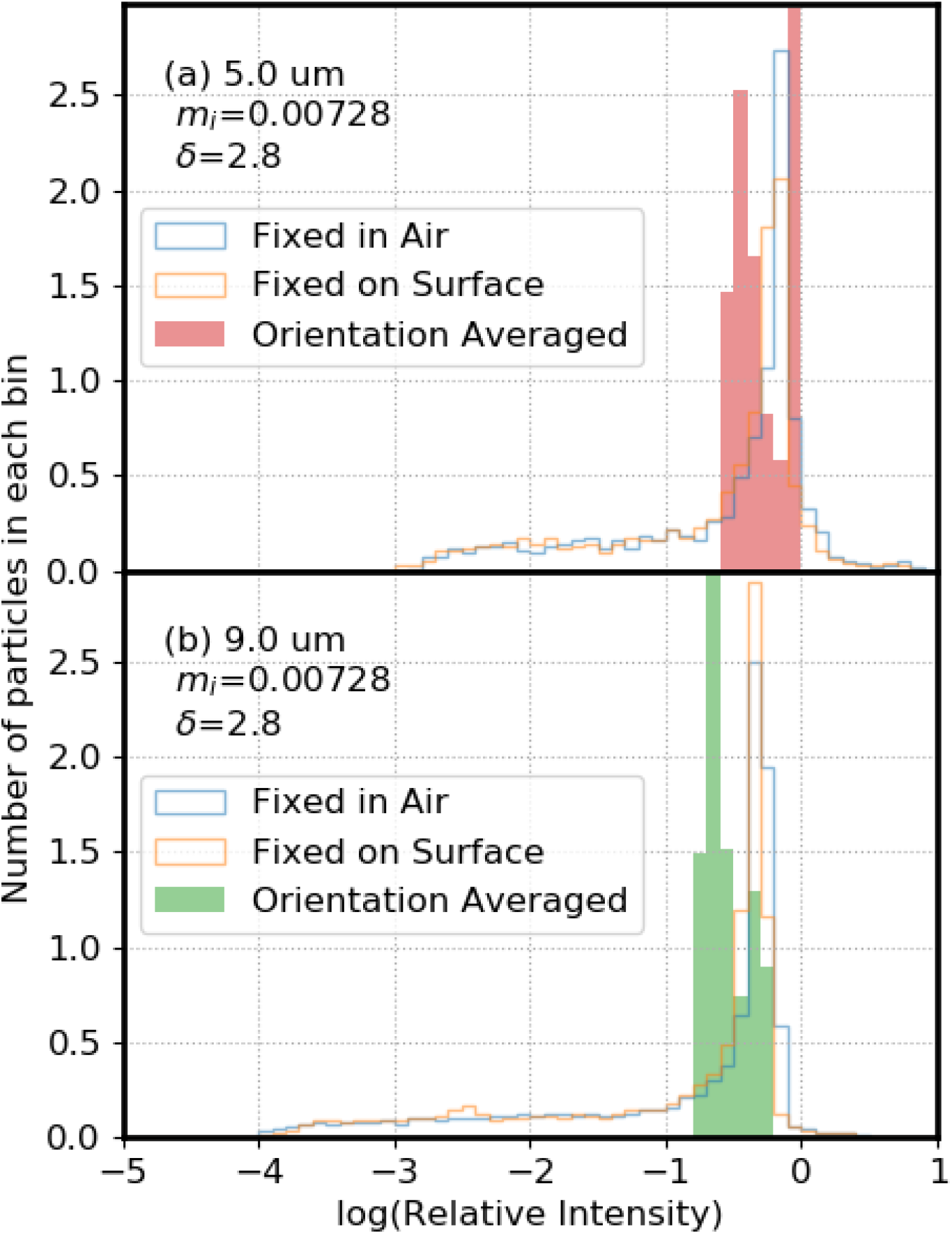
Relative numbers of virions in bins with the log(relative UV intensities) shown. The histograms in solid bars are for intensities averaged over all orientations. Histograms indicated by the blue line are for the same particle, in air, but in a single orientation. Histograms shown by the orange line are for the same particle, but held on a surface (m_r_ = 1.4) at a single orientation. In (a) the particle is 5-µm diameter. In (b) the particle is 9-µm diameter. The orientation-averaged simulations used in (a) 3000 virions (30 runs of 100 virions for a 9-µm particle), and in (b) 1000 particles (40 runs of 25 virions for a 5-µm particle). The area under the histograms has been normalized to 1.

In most situations where UV light is used to inactivate viruses in air the airflows are turbulent. Particle orientation with respect to a UV light source can vary by “spinning” (i.e., rotating about a particle axis), or by “tumbling” (i.e., rotation of the axis of the particle), or by following the airflow. Particles undergo Brownian translational and rotational diffusion (Fuchs, 1964) even in a small closed container. The extent to which the UV in a particle is equivalent to the orientation-averaged results depends upon the airflows (including turbulence), Brownian rotational diffusion, particle size, time of exposure, and the position(s) and spatial extent(s) of sources of UV light. For a sufficiently short illumination time and slow enough particle rotation, particle rotation during illumination is negligible and the particle is essentially illuminated from one angle, as if it were “fixed in air”.

In the cases of the particles averaged over all orientations, no particles 5- or 9-μm have *R*_*i*_ < 0.1. In contrast, the fixed-orientation 5-μm particles have 8.6 and 10% of virions with *R*_*i*_ < 0.01, and the 9-μm particles have 16.6 and 17.5% of virions with *R*_*i*_ < 0.01 and have 6.7 and 6.4% of virions with *R*_*i*_ < 0.001 (Table 3). That is, for an airborne UV-illuminated 9-μm particle with the properties used here, which rotates such that it is approximately illuminated uniformly by UV light from all directions, the best shielded virions are still exposed to a UV intensity approximately 1/7^th^ of the UV intensity a clean individual virion would be exposed to. If that same 9-μm particle were held on a surface, or illuminated for such a short time it could change orientation only a little, then approximately 17% of the virions are shielded so that their exposure to UV is 100 times less than is the single clean virion.

## 4. Discussion

### 4.1 Virions in Particles Can Be Partially Shielded from UV Light

#### 4.1.1 Particles on a Surface: UV Illumination from One Direction

a. Shielding of virions can be very significant even in weakly absorbing particles. Even when the penetration depth is 6.7 times greater than the particle diameter (Fig. 3a), 3.5% of the particles have *R*_*i*_ < 0.01 and 22.8% have *R*_*i*_ < 0.1. At such large ratios of δ/d the shielding is very largely refractive.
b. The effect of increased absorbance of the particle is apparent in Fig 3. As the penetration depth of the dried respiratory fluids decreases, the percent of *R*_*i*_ < 0.001 increases from 0.0 to 0.06, 6.4 and 11.8 for penetration depths of 59.9, 10.0, 2.8, and 1.4 µm, respectively.
c. Shielding of virions from UV light occurs with both the germicidal (260 nm) and solar (302 nm) light used, and the 5- and 9-µm diameter particles shown. Shielding from UV light is far less important in 1-µm diameter particles.
d. The effect of increased reflectance of the surface on shielding from UV appears to be small for the 5- and 9-µm diameter particles and two surfaces shown in Fig. 7 (air with *m*_*r*_ = 1.0, or a material with *m*_*r*_ = 1.4). However, reflective coatings on the walls of a room have been demonstrated to reduce times required for UV decontamination (Rutala et al. 2013; Krishnamoorthy and Tande, 2016). Such coatings can increase the UV intensity throughout the room and thus provide UV illumination from more directions and a higher total illumination intensity.

#### 4.1.2 UV Illumination from Multiple Directions Can Reduce Shielding from UV

UV intensities within refractively shielded regions can be increased by illuminating the particles from widely separated angles (Fig. 6 e-g), in cases where such illumination is possible. Wide-angle illumination can be achieved to varying degrees by using multiple UV light sources, and/or using movable or robotically controlled sources. Equal magnitude waves propagating in opposite directions have been used previously to increase the light intensities in the lowest regions of a sphere (Hill et al., 1998). Painting the walls and other surfaces with a highly UV-reflective paint (Ryan et al., 2010; Rutala et al., 2013; Krishnamoorthy and Tande, 2016) also typically increases the UV illumination from many angles.

#### 4.1.3 Particles in Air May Rotate to be UV Illuminated from Many Directions

Refractive shielding of virions in particles that rotate through all orientations is small, as illustrated in Fig. 7. Absorptive shielding can still be significant, but did not have a large effect with the *m*_*i*_ used for airway fluids even in 9-µm particles. Whether or not an airborne particle of a certain size rotates through enough angles to have a only small amount of refractive shielding and a smaller effect of absorptive shielding depends on the particle size and shape, the illumination times, positions of the UV lights, and airflows (turbulent, laminar, circuitous or straight). If the UVGI system is in a flow-through configuration (such as a HVAC system), larger particles may be removed by filters at the entrance of the UVGI system.

### 4.2 Uncertainties and potential effects on the findings

#### 4.2.1 Variations and uncertainties in compositions and complex refractive indexes

Several problems arise in modeling the UV within virus-containing particles even when focusing primarily on virions in particles in dried respiratory droplets. Large variations occur in concentrations in airway fluids of materials such as mucins, other proteins, DNA, inorganic salts, uric acid, and other components of broken cells (Wyllie et al., 2020; Hawkins et al., 1963; Araujo et al., 2020). Even for a single person these concentrations may depend upon hydration, exercise, odors, time of day, time since last meal, etc. They also vary from person to person. The uncertainty associated with unknown concentrations and absorptivities of the primary absorbing molecules arises in the common case where a large predominance of non-viral material in a particle (e.g., lysed cell material) is not understood. To account for such variability and see which parameters the shielding most depends upon, the model results are calculated for the m_*i*_ spanning a large range. Variations and uncertainties in the densities and compositions of the dried particles may be the largest sources of differences between the calculations illustrated here and any specific particle of dried respiratory fluids. The absorptive and refractive shielding illustrated here are general; however, variations in measured saliva and other fluids are large. For example, nucleic acids are some of the most, if not the most, important UV absorbing materials in saliva at 260 nm. However, reported concentrations of DNA in saliva vary widely from 0.01 to 0.28 mg/ml as measured by Poehls et al., (2018). Review of the literature revealed no measurements of RNA concentrations in saliva other than measurements of RNA viruses. The particles here were assumed to be very dry, as in low humidity environments. Higher humidity would result in higher water contents of the particles, lower densities and larger penetration depths.

#### 4.2.2 Dried droplets are neither perfectly spherical nor optically homogeneous

Actual dried particles of airway fluids are not totally spherical and not likely to be optically homogeneous. For particles formed by drying droplets of simulated respiratory fluids (water, mucin, NaCl and DPPC) on a superhydrophobic surface, the particles with sizes similar to those modeled here were somewhat spherical. In the larger dried droplets, crystals of mucin, NaCl, and DPPC formed, in some cases making the particles inhomogeneous and less spherical (Vejerano and Marr, 2018). The use of four components (NaCl, mucin, DPPC and water) for the simulated saliva may be the reason for this crystallization into what appear to be somewhat pure materials, and the resulting nonsphericity. Human saliva, on the other hand, is highly complex (Edgar et al., 2012; Poehls et al., 2018; Hawkins et al. 1963; Araujo et al., 2020; Peden et al., 1990) as seen in Section 2.2 above. There are many different inorganic (Na^+^, K^+^, Mg^2+^, Ca^2+^, Cl^-^, PO_4_^3-^, HCO_3_^-^, SCN^-^, F^-^) electrolytes, organic acids (lactic and uric acids), glucose, amino acids, peptides, proteins, nucleic acids, etc. Also, DNA and RNA in saliva increase the viscosity. It is therefore not clear which materials, if any, crystalize as pure components when droplets of airway fluids dry, and therefore, whether this possible cause of nonsphericity and inhomogeneity is relevant in dried droplets of actual respiratory fluids.

Although there are uncertainties about the shapes and homogeneities of dried droplets of airway fluids, some general comments about shielding in such particles can be made. First, as illustrated in the first panel of Fig. 1, neither refractive nor absorptive shielding requires spherical particles. Second, given the complexity of airway fluids, it seems unlikely that differences in refractive index within the particle will be as large as those between pure NaCl and a typical protein; we think the maximum differences are likely to be far less. Also, a small amount of inhomogeneity appears to have a small effect. For example, adding 90 randomly located virions to a 9 µm particle containing 10 virions resulted in changes in *R*_*i*_ of the initial 10 virions ranging from 0.03% to -0.27%. Third, if the surface of a particle were sufficiently rough that it scattered the UV illumination light in all directions as the light entered the particle, then, as can be envisioned using Fig. 1a and 1b, refractive shielding could far less than it would be in an otherwise identical particle with a smooth surface. Fourth, inhomogenities such as large air pockets forming within droplets as they dry, or large cracks occurring as the particles shrink, may have large impacts on the UV intensity distribution. It is, however, difficult to see how a single air pocket or crack would eliminate the potential for refractive shielding, at least at some illumination angles, unless there were multiple cracks and the sides of them were rough. Also, air pockets or wide cracks could decrease absorptive shielding for some illumination angles. Fifth, the locations of the virions in particles are not known (Vejerano and Marr, 2018). However, as a droplet dries, and as water evaporates and leaves behind a more concentrated region near the droplet surface, the 100-nm virions are likely to diffuse more slowly toward center of the droplet (where the concentration of everything except water is lower) than do proteins and smaller molecules. If that is the case, and so if virions are more likely to occur in the outer fraction of the dried droplet, then the actual fraction of particles which can be refractively shielded is likely to be more than is now indicated in the figures and Table 3. Figure 2 shows that the most shielded virions, for a particle on a surface and a normally incident plane wave, are near the outside of the particle. Even for *δ* = 2.84 µm with the 9-µm particle in Fig 2b, particles within approximately 2 µm of the center point of the particle appear to have *R*_*i*_ > 0.1. In the outer approximately 1.5 µm of the hemisphere closest to the surface some particles are well-shielded. Thus, if the virions localize to the surface of the dried droplet as hypothesized above, the overall shielding would increase for fixed particles on a surface.

### 4.3 Interventions to Circumvent the Effects of Shielding of Virions from UV Light

For particles on surfaces, using multiple UV-light sources positioned to illuminate from widely varying angles can strongly reduce shielding of virions (see Fig 6 e-g and Table 3). Similar reductions in shielding can also be obtained using movable light sources. Painting the walls and other surfaces in a room with highly reflective paint can increase the UV intensities in otherwise shielded regions, partly by increasing the number of directions from which a particle is illuminated, and can increase the total intensity illuminating the particles.

For particles in air, UVGI systems may be designed so that particles are illuminated from approximately all orientations, even for large particles moving through the system too quickly to rotate through a large angular range. Or the system may be designed so that all particles have time to rotate though a sufficiently large range of angles to approximate the all orientations results of Fig. 7. In flow-through UVGI systems, filters which remove a high percentage of particles larger than, e.g., 5 µm, are common (Kowalski, 2009). Such filters reduce the need to design the UV optics to inactivate larger particles. Rotation of particles in turbulent air flowing through a system (UVGI or other) is a topic that appears to need further investigation.

## 5. Conclusions

Virions in particles may be partially shielded from UV. Refractive shielding is a primary means of protection from UV, and occurs even in weakly UV absorbing particles. It results primarily from refraction and reflection of light at the front surface of the particle. Absorptive shielding is most important at the wavelengths of germicidal UV (260 nm used here), where nucleic acids and aromatic amino acids absorb UV light well, especially in large particles with diameters several times larger than the penetration depth. Absorptive shielding is much less important at the solar UV wavelength (302 nm) used here.

The extent to which these modeling results apply to actual dried droplets of respiratory fluids depends on the characteristics of the actual dried particles. In this study the optical properties of the droplets are assumed to be homogeneous, except for the virions, and all droplets and virions are assumed to be spherical. The effects of nonsphericity, inhomogeneous refractive indexes, and the effects of location of the virions are discussed in Section 4.2. As illustrated in Fig. 1a, and readily envisioned from the ray optics (Chowdhury et al., 1991) shielding can occur in nonspherical particles which have a smooth surface. The nonsphericity that could strongly decrease shielding would be a rough surface that scattered the UV light into the particle in all directions.

The key point of relevance to designers and users of UVGI systems is: these results suggest that shielding of virions from UV can be reduced by illuminating virion-containing particles with light from widely separated directions (see Figs. 6e-g).

For inactivation of virions in particles on surfaces, illuminating with UV from more, widely separated, directions can help reduce the refractive shielding (Figs. 6e-g). Painting walls and other surfaces in a room with a highly UV-reflective paint (Rutala et al., 2013; Krishnamoorthy and Tande, 2016) can help increase the angles from which the UV illuminates the particles on surfaces, and increase the UV intensity in the room, as compared with the same room with less reflective surfaces.

For inactivation of virions in particles in air, UV illumination of particles from many angles can be easier to achieve because particles change their orientation with respect to the source(s) of UV illumination as they move through a UVGI system and rotate to some degree. Do the particles of the relevant sizes rotate sufficiently as they traverse the UVGI system so that shielding of virions from UV is low? For a 5- or 9-µm diameter particle traversing a UVGI system in 1 s, the Brownian rotational diffusion is too small to avoid shielding from UV if a single UV source is employed and reflectivities of surfaces are small. Turbulent airflows may provide adequate rotational motion for the virions in particles to avoid UV shielding in large particles. To reduce the problem of needing rotation to avoid shielding, UVGI systems that illuminate particles somewhat uniformly from all directions as they flow through the system may be ideal. Alternatively, UVGI systems which illuminate from one direction, or directions separated by a not-large angle, may reduce shielding adequately if they are designed so that even the largest particles have sufficient time to rotate through a sufficiently wide range of angles as they flow through the system. Or, in many cases, the large particles may be removed from the airflow with a suitable filter.

## Data Availability

Data available upon request.

## Code Availability

Researchers interested in the MSTM codes to calculate the results can email D.W. Mackowski. Those interested in reproducing the calculations, figures or Table 3 can email D. C. Doughty. Those interested in reproducing Tables 1 or 2 can email S. C. Hill.

## Acknowledgment

The authors acknowledge partial support from the Defense Threat Reduction Agency (DTRA). Hill and Doughty acknowledge funding from the US Army DEVCOM Army Research Laboratory.

